# Sex difference in the associations of socioeconomic status, cognitive function and brain volume with dementia in old adults: Findings from the OASIS study

**DOI:** 10.1101/2023.01.05.23284240

**Authors:** Sophia Z Liu, Ghazaal Tahmasebi, Ying Sheng, Ivo D Dinov, Dennis Tsilimingras, Xuefeng Liu

## Abstract

**Background:** Sex differences in the association of cognitive function and imaging measures with dementia have not been fully investigated while sex-based investigation of dementia has been discussed. Understanding sex differences in the dementia-related socioeconomic, cognitive, and imaging measurements is important for uncovering sex-related pathways to dementia and facilitating early diagnosis, family planning, and cost control.

**Methods:** We selected data from the Open Access Series of Imaging Studies with longitudinal measurements of brain volumes on 150 individuals aged 60 to 96 years. Dementia status was determined using the Clinical Dementia Rating (CDR) scale, and Alzheimer’s disease was diagnosed as a CDR of ≥ 0.5. Generalized estimating equation models were used to estimate the associations of socioeconomic, cognitive and imaging factors with dementia in men and women.

**Results:** Lower education affected dementia more in women than in men. Age, education, Mini-Mental State Examination (MMSE), and normalized whole-brain volume (nWBV) were associated with dementia in women whereas only MMSE and nWBV were associated with dementia in men. Lower socioeconomic status was associated with a reduced estimated total intracranial volume in men, but not in women. Ageing and lower MMSE scores were associated with reduced nWBV in both men and women.

**Conclusions:** The association between education and prevalence of dementia differs in men and women. Women may have more risk factors for dementia than men.

## INTRODUCTION

Dementia refers to a group of symptoms associated with a decline in mental ability characterized by memory, reasoning, and other thinking skills.^1^ Five million US adults of more than 65 years of age had dementia in 2014, and it is projected to be nearly 14 million by 2060.^2^ Worldwide, approximately 50 million people live with dementia and this number is expected to triple by 2050 with the aging of the population, presenting a substantial challenge to patients, families, and society.^3^

Alzheimer’s disease (AD) is the most common cause of dementia accounting for 60-80% of dementia cases.^4^ The common risk factors for AD dementia include age, genetics, family history, race/ethnicity,^5^ low education, excessive alcohol consumption, smoking, obesity, hypertension, hearing impairment, depression, physical inactivity, diabetes, poor heart health, and traumatic brain injury.^6, 7^ Addressing preventable risk factors (e.g. low education and low socioeconomic status) may reduce the risk of cognitive decline and prevent or delay up to 40% of dementia cases.^7^

Sex-based investigation of dementia has been discussed and implemented.^8, 9^ Although education and socioeconomic status (SES) have been associated with dementia with lower education level and lower SES groups presenting a higher risk for dementia,^10-12^ little information is known about how the associations differ in men and women. Moreover, the diagnosis of dementia may involve screening medical tests (e.g. MMSE, Mini-Mental State Examination) for measuring cognitive impairment and magnetic resonance imaging (MRI) for measuring brain volumes. Sex differences in the association of these measures with dementia have not been fully investigated. Understanding sex differences in dementia-related socioeconomic, cognitive, and MRI-imaging measurements will uncover the sex differences in the pathway to dementia and facilitate early diagnosis, family planning, and cost control.

The aim of this study is to evaluate sex differences in education, cognitive function, and brain volumes with the ultimate goal of determining whether there is a difference in the associations of these factors with the prevalence of dementia in men and women.

## METHODS

### Data Source and Subjects

The dataset was obtained from the Open Access Series of Imaging Studies (OASIS II) project with longitudinal MRI data on 150 individuals aged 60 to 96 years.^13^ Subjects in the OASIS II were selected from a larger longitudinal pool of individuals who had participated in MRI studies at the Washington University Alzheimer Disease Research Center (ADRC). The selection was based on the availability of at least two separate visits in which clinical and MRI data were collected, at least three acquired T1-weighted images per imaging session, and right-hand dominance.^13, 14^ The ADRC’s normal and cognitively impaired subjects were recruited primarily through media appeals and word of mouth, with 80% of subjects initiating contact with the center and the remainder being referred by physicians. All subjects participated in accordance with guidelines of the Washington University Human Studies Committee. Approval for public sharing of the anonymized data was also specifically obtained. The OASIS II included 65 individuals with very mild to moderate AD at their initial visit, and 14 individuals characterized as nondemented at the time of one or more scans and then clinically determined to have AD at the time of a subsequent scan. Data were acquired on the same scanner using identical procedures.

All subjects were screened for inclusion, and each subject underwent the ADRC’s full clinical assessment as described below. Subjects with a primary cause of dementia other than AD (e.g., vascular dementia and primary progressive aphasia) were excluded, as were subjects with gross anatomical abnormalities evident in their MRI images (e.g. large lesions and tumors). However, subjects with age-typical brain changes (e.g. mild atrophy and leukoaraiosis) were included. MRI acquisitions were typically obtained within one year before or after the clinical assessment of each subject. Each subject was scanned on two or more separate occasions with an average delay of 719 days between the visits. The final data set included 150 subjects and 373 imaging sessions.

### Clinical Assessment

A Mini-Mental State Examination (MMSE)^15^ including 11 questions was conducted by trained physicians to assess the cognitive function in six areas of mental abilities, including orientation to time and place, attention and concentration, short-term memory (recall), language skills, visuospatial abilities, and ability to understand and follow instructions. The MMSE with a 30-point score was used to screen patients for cognitive impairment (A score of 24 or lower) and track changes in cognitive functioning over time in the study.

Dementia status was established and staged using the Clinical Dementia Rating (CDR) scale.^16^ The CDR is a dementia staging instrument that rates subjects for impairment in each of six domains: memory, orientation, judgment and problem solving, function in community affairs, home and hobbies, and personal care. Based on the collateral source and subject interviews, a global CDR score was derived from the individual ratings in each domain. A global CDR of 0 indicates no dementia, and CDR of 0.5, 1, 2 and 3 represent very mild, mild, moderate, and severe dementia, respectively. The clinical diagnosis of AD was determined as a CDR ≥0.5.

### Image Acquisition and Processing

As detailed in the previous publications,^13, 17^ each subject had 3 to 4 T1-weighted magnetization prepared rapid gradient echo (MP-RAGE) images acquired on a 1.5-Tesla Vision scanner (Siemens, Erlangen, Germany) in a single imaging session. The individual scan files were converted from Siemens proprietary IMA format into 16-bit NiFTI1 format using a custom conversion program. The images were then corrected for inter-scan head movement and spatially warped into the atlas space using a rigid transformation that differed in process from the original piecewise scaling. The resulting transformation places the brains in the same coordinate system and bounding box as in the original atlas. Given the age range of the present sample, an old-only atlas target was employed. For registration, a 12-parameter affine transformation was computed to minimize the variance between the first MP-RAGE image and the atlas target. The remaining MP-RAGE images were registered to the first (in-plane stretch allowed) and resampled by transforming the composition into a 1-mm isotropic image in the atlas space. The result was a single, high-contrast, averaged MP-RAGE image in the atlas space. Subsequent steps included skull removal by the application of a loose-fitting atlas mask and correction for intensity inhomogeneity due to non-uniformity in the magnetic field. Intensity variation was corrected across contiguous regions, based on a quadratic inhomogeneity model fitted to data from a phantom.

### Estimated Total Intracranial Volume and Normalized Whole-brain Volume

The procedures used for measuring intracranial and whole-brain volumes have been described previously.^13, 17, 18^ The estimated total intracranial volume (eTIV) was computed by scaling the manually-measured intracranial volume of the atlas by the determinant of the affine transform connecting each individual’s brain to the atlas. This method is minimally biased by atrophy and is proportional to manually measured total intracranial volume.

Normalized whole-brain volume (nWBV) was computed using the FMRIB Automated Segmentation Tool in the FSL software suite (https://fsl.fmrib.ox.ac.uk/fsl/fslwiki) which was developed for the segmentation of trained tissues. The image was first segmented to classify the brain tissue as cerebrospinal fluid (CSF), gray matter, or white matter. The segmentation procedure iteratively assigned voxels to tissue classes based on the maximum likelihood estimates of a hidden Markov random field model using an Expectation-Maximization Algorithm. The model used spatial proximity to constrain the probability that voxels of a given intensity are assigned to each tissue class. Finally, the nWBV was computed as the proportion of all voxels within the brain mask classified as tissue (either gray or white matter). The unit of normalized volume is percentage, which represents the percentage of the total white and gray matter voxels within the estimated total intracranial volume.^18^

### Other Characteristics

Socioeconomic characteristics of the subjects were collected, including age in years at the time of image acquisition, sex (men and women), years of education, and socioeconomic status (SES). Education was classified into two groups: High school or below and college or above in terms of years in school. SES was assessed using the Hollingshead Index of Social Position^19^ and classified into categories ranging from 1 (highest status) to 5 (lowest status).

### Statistical Analysis

Sample characteristics were summarized as means (standard errors) for continuous variables and counts (percentages) for categorical variables. Independent t tests were used to test the significance of differences in means, and chi-square tests were used to test the significance of differences in percentages across sex and dementia groups. Generalized estimating equation (GEE) models were conducted to estimate adjusted odds ratios (ORs) with 95% confidence intervals (CIs) that measure the associations (main effects) of socioeconomic characteristics with eTIV and nWBV, and the associations of eTIV and nWBV with dementia. The differences in the associations between men and women (interaction effects) were examined using GEE models by adding the first-order interaction terms of sex and other characteristics. Chi-square tests were used to test the significance of the associations, including the main and interaction effects in the GEE models. Stratification analyses by sex were also conducted to identify the significant characteristics associated with eTIV, nWBV and dementia in men and women, separately. In this study, an association estimate was considered statistically significant when the corresponding p-value was less than 0.05. All data analyses were performed using SAS 9.4 (Cary, NC: SAS Institute Inc.).

## RESULTS

The present study sample consisted of 88 women (58.7%) and 62 men (41.3%) aged 60 to 96 years (Table 1). At the time of their initial visit, the average age of the subjects was 75.4 years, and 65 (42.7%) had dementia with a CDR score greater than 0. Of all the subjects, 35.3% received a high school education or below. Compared to men, women had lower eTIV (1390.9 vs. 1593.0, p<.0001), higher MMSE (28.1 vs. 26.8, p=.0073), higher nWBV (74.3 vs. 72.6, p=.0029), and lower dementia rate (32.9% vs. 58.1%, p=.0022). Subjects with dementia (vs. non-dementia) had fewer women (44.6% vs 69.4%, p=.0022), received more high school education or below (50.8% vs. 23.5%, p=.0005), poorer SES (2.7 vs 2.3, p=.034), lower MMSE (25.4 vs. 29.2, p<.0001), and lower nWBV (72.5 vs. 74.5, p=.0008).

**Table 1.**
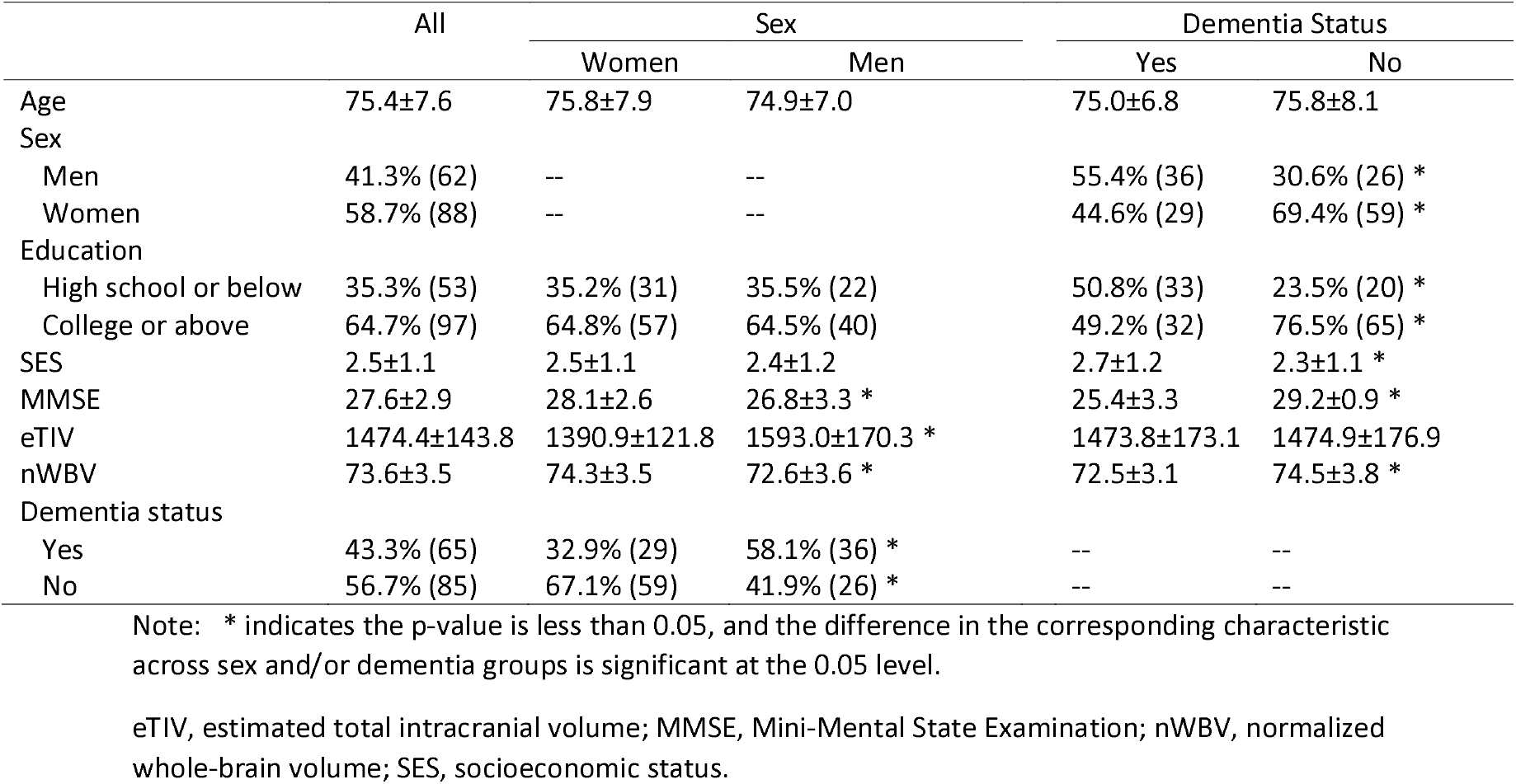
Sample characteristics by sex and dementia status (n=150)

|  | All | Sex |  | Dementia Status |  |
| --- | --- | --- | --- | --- | --- |
|  |  | Women | Men | Yes | No |
| Age | 75.4±7.6 | 75.8±7.9 | 74.9±7.0 | 75.0±6.8 | 75.8±8.1 |
| Sex |  |  |  |  |  |
| Men | 41.3% (62) | -- | -- | 55.4% (36) | 30.6% (26) * |
| Women | 58.7% (88) | -- | -- | 44.6% (29) | 69.4% (59) * |
| Education |  |  |  |  |  |
| High school or below | 35.3% (53) | 35.2% (31) | 35.5% (22) | 50.8% (33) | 23.5% (20) * |
| College or above | 64.7% (97) | 64.8% (57) | 64.5% (40) | 49.2% (32) | 76.5% (65) * |
| SES | 2.5±1.1 | 2.5±1.1 | 2.4±1.2 | 2.7±1.2 | 2.3±1.1 * |
| MMSE | 27.6±2.9 | 28.1±2.6 | 26.8±3.3 * | 25.4±3.3 | 29.2±0.9 * |
| eTIV | 1474.4±143.8 | 1390.9±121.8 | 1593.0±170.3 * | 1473.8±173.1 | 1474.9±176.9 |
| nWBV | 73.6±3.5 | 74.3±3.5 | 72.6±3.6 * | 72.5±3.1 | 74.5±3.8 * |
| Dementia status |  |  |  |  |  |
| Yes | 43.3% (65) | 32.9% (29) | 58.1% (36) * | -- | -- |
| No | 56.7% (85) | 67.1% (59) | 41.9% (26) * | -- | -- |

Note: \* indicates the p-value is less than 0.05, and the difference in the corresponding characteristic across sex and/or dementia groups is significant at the 0.05 level.
eTIV, estimated total intracranial volume; MMSE, Mini-Mental State Examination; nWBV, normalized whole-brain volume; SES, socioeconomic status.

The first-order interaction analysis of sex and other characteristics showed a significant sex-by-SES interaction effect on eTIV (p=.0064), indicating that the associations of SES with eTIV differ in men and women. No interaction effects of sex and other characteristics on nWBV were found. Stratification analysis by sex (Table 2) showed that lower SES was associated with reduced eTIV in men (−59.24±25.17, p=.02), but not in women. Older age was associated with reduced nWBV (−0.33±0.04, p<.0001 for men; -0.33±0.02, p<.0001 for women), and higher MMSE scores were associated with increased nWBV (0.15±0.02, p<.0001 for men; 0.16±0.06, p=.006 for women) in both men and women.

**Table 2.** GEE models for associations of socioeconomic characteristics with eTIV and nWBV by sex

|  | Women |  |  |  |  |  | Men |  |  |  |  |  |
| --- | --- | --- | --- | --- | --- | --- | --- | --- | --- | --- | --- | --- |
|  | eTIV |  |  | nWBV |  |  | eTIV |  |  | nWBV |  |  |
|  | Estimates | Standard Errors | P values | Estimates | Standard Errors | P values | Estimates | Standard Errors | P values | Estimates | Standard Errors | P values |
| Age | 0.90 | 0.90 | 0.32 | -0.33 | 0.02 | <.0001 | 2.93 | 1.67 | 0.08 | -0.33 | 0.04 | <.0001 |
| Education (High school or below vs. College or above) | 7.94 | 31.81 | 0.80 | -1.61 | 0.85 | 0.06 | -18.38 | 65.54 | 0.78 | 0.07 | 1.38 | 0.96 |
| SES | -15.60 | 13.62 | 0.25 | 0.46 | 0.38 | 0.22 | -59.24 | 25.17 | 0.02 | 0.28 | 0.54 | 0.61 |
| MMSE | -2.32 | 1.17 | 0.05 | 0.16 | 0.06 | 0.006 | 0.29 | 1.65 | 0.86 | 0.15 | 0.02 | <.0001 |

Notes: eTIV, estimated total intracranial volume; MMSE, Mini-Mental State Exam; nWBV, normalized whole brain volume; SES, socioeconomic status.

The changes in eTIV and nWBV with age were presented in Figure 1. The figure showed that nWBV decreased with age while eTIV did not change with age, with parallel trend lines (approximately identical slopes) in men and women indicating that there were no sex differences in the associations of age with eTIV and nWBV.

**Figure 1.**
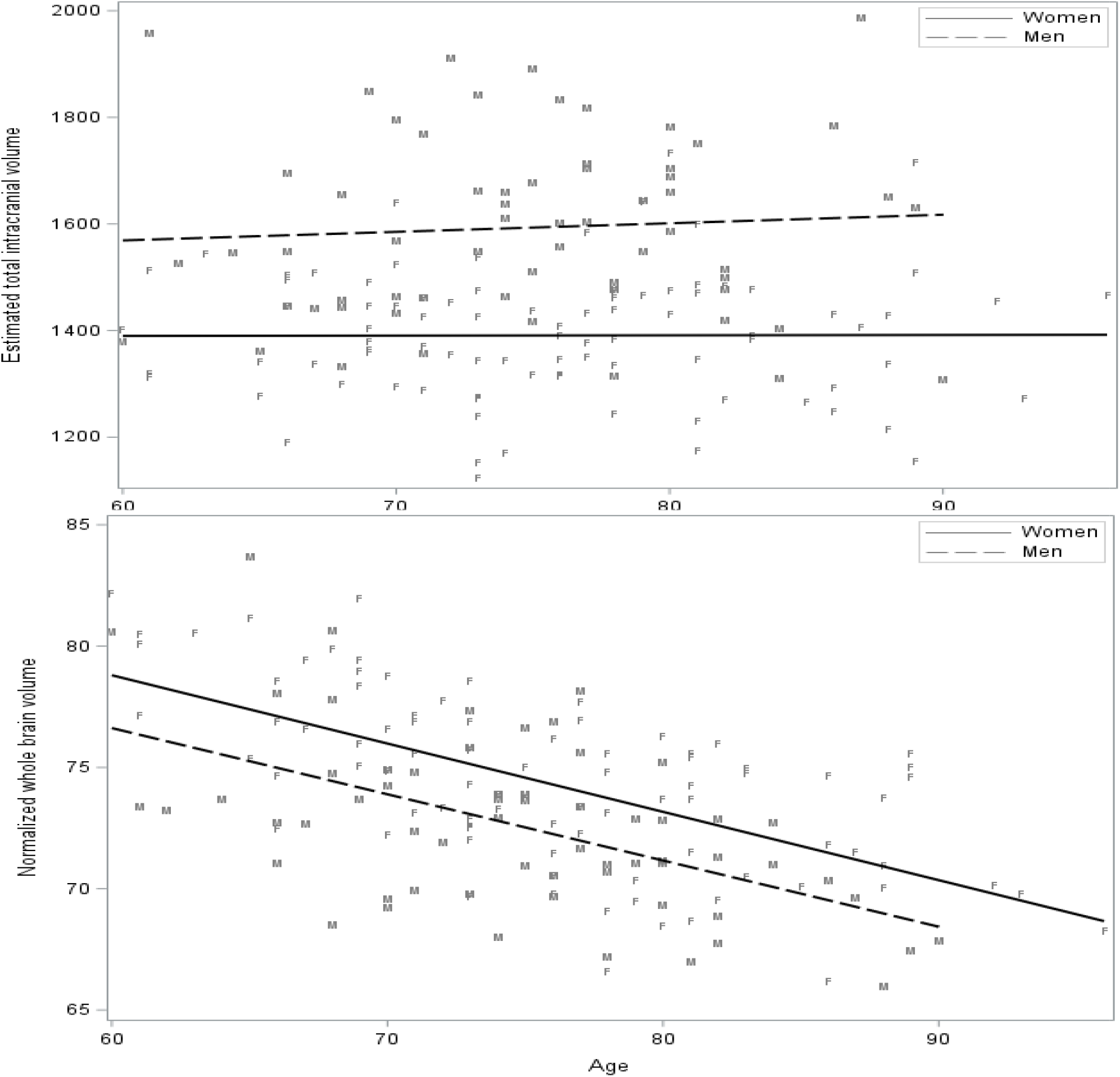
Plots of automated anatomical measures versus age. Each point represents a unique subject (F, female; M, male) from a single scanning session. (Top) Estimated total intracranial volume: The trend lines are separately drawn for females and males. (Bottom) Normalized whole brain volume: The trend lines are separately drawn for females and males.

In the general study sample, including all old subjects, age, sex, MMSE, eTIV, and nWBV were significant predictors of dementia (Figure 2). GEE models involving interactions showed that the interaction effect of sex and education on dementia was significant, indicating a difference in the association of education with dementia in men and women. Stratification analysis by sex revealed that age, education, MMSE score, and nWBV were associated with dementia in women; however, only MMSE and nWBV were associated with dementia in men (Table 3). Lower education level was associated with a higher risk of dementia in women but not in men.

**Table 3.**
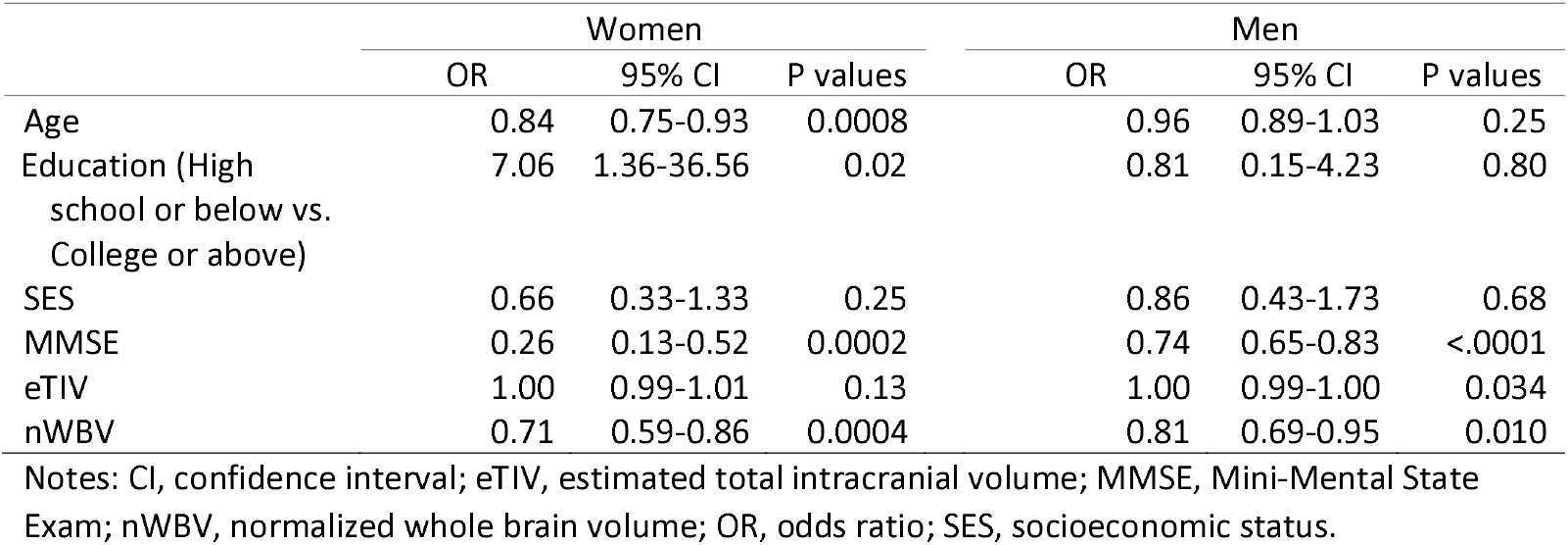
GEE models for associations of socioeconomic and brain imaging characteristics with dementia by sex in the longitudinal study sample

**Figure 2.**
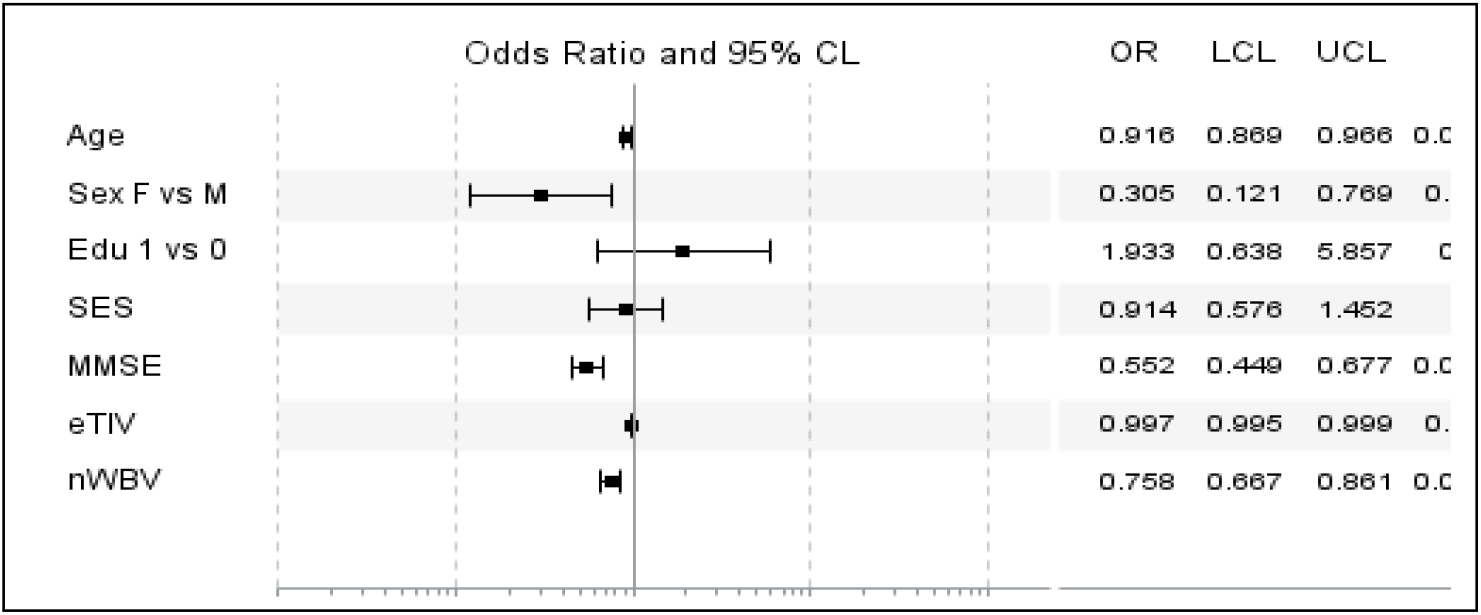
Odds ratio plot of dementia related to the socioeconomic and imaging characteristics. The plot shows the odds ratios of dementia with 95% confidence limits and p values. eTIV, estimated total intracranial volume; LCL, lower confidence limit; MMSE, Mini-Mental State Exam; nWBV, normalized whole brain volume; OR, odds ratio; SES, socioeconomic status; UCL, upper confidence limit.

## DISCUSSION

In this study, we investigated older participants aged 60-96 years and demonstrated that the prevalence of dementia was lower in women than in men, and nWBV and MMSE were larger and eTIV was smaller in women. Aging and MMSE scores were associated with the changes in nWBV in both men and women with older age and lower MMSE scores indicating a decline in brain volume, while there was no sex difference in this relationship. Lower SES status was associated with smaller eTIV; however this association was stronger in men. The significant predictors of dementia were age, education, MMSE and nWBV in women, and MMSE and nWBV in men with common predictors of MMSE and nWBV for both sexes. Lower education level had a stronger association with dementia in women than in men.

Based on the frequency count of all individuals with AD, more women than men are living with a diagnosis of AD.^4, 8, 20^ Women also have a greater lifetime risk of developing AD.^4^ An important contributor to this sex difference in both the frequency and the lifetime risk is that women live longer than men. However, when assessing the prevalence/rate (total number with disease divided by the total number of individuals in the population) of AD by sex, the results are conflicting. Several studies in Europe have reported a higher prevalence of AD among women.^21^ However, the studies in the United States,^22^ and around the world,^23^ have not reported a significant sex difference. In the present study sample aged 60-96 years, we found that the dementia rate was higher in women than in men, which is consistent with the previous reports.^21, 24, 25^ The conflicting results appear to be somewhat dependent on the time period and geographic regions where the studies were conducted.

nWBV has been reported to decline with age, while eTIV does not vary much with age.^17, 26^ Our results confirm those of previous reports and further show that the trend in nWBV and eTIV over age did not differ in men and women, although baseline nWBV was higher and basline eTIV was lower in women than in men (Figure 1 and Table 1). A previous study demonstrated the presence of significant differences in neuroanatomical structure volumes between men and women in a population sample of participants above 66 years of age.^27^ It revealed that the adjusted gray and whiter matter volume was larger in women. Since the total brain volume is calculated as the sum of the gray and white matter volumes, this finding is consistent with our analysis that average nWBV was higher in women than in men. Our GEE analysis showed that nWBV was negatively linked to the prevalence of dementia with larger nWBV, indicating a lower risk of dementia. The results together could partly explain why the dementia rate was lower in women in the present study sample, while the strength of the association of nWBV with dementia did not differ in men and women.

Our analysis showed that after adjusting for socioeconomic factors, MMSE and nWBV were negatively associated with dementia in both men and women while there was not a sex difference in this association. The MMSE has been used to screen patients for cognitive impairment, track changes in cognitive functioning over time, and often to assess the effects of therapeutic agents on cognitive function. Its utility decreases when patients with mild cognitive decline and psychiatric conditions are assessed,^28, 29^ and should not be used alone as a tool for diagnosing dementia.^30^ As a similar strength of its association with dementia was found in both sex, higher average MMSE in women may be the contributor to the sex difference in the prevalence of dementia. nWBV is the normalized total brain volume corrected for the intracranial volume. Consistent with this study, it has been shown to be larger in women than in men, although uncorrected brain volume measurements, such as gray matter and white matter are smaller in women.^27^ As the strength of its association with dementia is the same in both men and women, similar to MMSE, the higher average nWBV in women may be the contributor to lower dementia rate in the female group. In addition, MMSE score and nWBV both declined with age. The role of MMSE and nWBV in relation to dementia may follow the pathway: Aging factors→decline in MMSE and nWBV→dementia In this study, lower SES was found to be associated with smaller eTIV, and the association of SES with eTIV was weaker in women than in men. eTIV appeared to be negatively associated with dementia, yet compared to MMSE and nWBV, its effect size was smaller in terms of odds ratios (very close to 1, indicating no effect) that characterize the risk of the disease. The eTIV did not change with age (Figure 1), indicating that aging-related factors may not affect eTIV. Whether the eTIV can predict the diagnosis of dementia requires further investigation.

Socioeconomic characteristics, including educational level, income level, and occupation, have been investigated as potential risk factors for dementia.^12, 31, 32^ A Denmark population study with 10,191 individuals reported that higher household income (vs. lower income) were associated with less likelihood of dementia diagnosis after referral, and led to earlier dementia diagnosis and less severe disease at diagnosis.^12^ Another study systematically reviewed 71 studies with 88 study populations and found that lower education is associated with a greater risk for dementia in many but not all studies.^31^ An epidemiologic project studied 10,781 Health and Retirement Study participants interviewed from 1998-2012 (US) and showed that compared to low SES at all 3 time points (childhood, early adulthood, and older adulthood), stable high SES predicted the best memory function and slowest decline, high school completion had the largest estimated effect on memory, and high late-life income had the largest estimated benefit for slowing memory declines. In the present study, we reviewed 150 participants aged 60-96 years and found that education and SES were not associated with dementia (Figure 2). However, the interaction and stratification analysis with GEE models showed that sex moderated the association between education and dementia, and lower education level was associated with a higher risk of dementia in women, but not in men. The level of education associated with the risk of dementia may vary with geographic regions and study populations and more years of education may not uniformly attenuate the risk. Sex differences in such associations and the underlying pathways need to be further investigated in the future research.

## Acknowledgement

We would like to express our thanks to the Washington University Alzheimer Disease Research Center for the collective and collaborative efforts to integrate neuroimaging datasets and make the Open Access Series of Imaging Studies (OASIS) project with longitudinal imaging measurements freely available to the scientific community. This effort provides a high-quality data source for training next-generation neuroscience and radiology researchers and facilitates future discoveries in basic and clinical neuroscience.

## Ethnical Statement

Public sharing of the anonymized data was approved by the Washington University Human Studies Committee.

## Data Availability

Analysis ata will be made available by the corresponding author to interested researchers upon request via email

